# Human-level information extraction from clinical reports with fine-tuned language models

**DOI:** 10.1101/2024.11.18.24317466

**Authors:** Longchao Liu, Long Lian, Yiyan Hao, Aidan Pace, Elaine Kim, Nour Homsi, Yash Pershad, Liheng Lai, Thomas Gracie, Ashwin Kishtagari, Peter R Carroll, Alexander G Bick, Anobel Y Odisho, Maggie Chung, Adam Yala

**Author notes:** **Corresponding author:** Adam Yala PhD, Address: 2177 Hearst Ave, Berkeley, CA 94709. Longchao Liu and Long Lian are joint first authors. Adam Yala and Maggie Chung are joint senior authors.

## Abstract

Extracting structured data from clinical notes remains a key bottleneck in clinical research. We hypothesized that with minimal computational and annotation resources, open-source large language models (LLMs) could create high-quality research databases. We developed Strata, a low-code library for leveraging LLMs for data extraction from clinical reports. Trained researchers labeled four datasets from prostate MRI, breast pathology, kidney pathology, and bone marrow (MDS) pathology reports. Using Strata, we evaluated open-source LLMs, including instruction-tuned, medicine-specific, reasoning-based, and LoRA-finetuned LLMs. We compared these models to zero-shot GPT-4 and a second human annotator. Our primary evaluation metric was exact match accuracy, which assesses if all variables for a report were extracted correctly. LoRa-finetuned Llama-3.1 8B achieved non-inferior performance to the second human annotator across all four datasets, with an average exact match accuracy of 90.0 ± 1.7. Fine-tuned Llama-3.1 outperformed all other open-source models, including DeepSeekR1-Distill-Llama and Llama-3-8B-UltraMedical, which obtained average exact match accuracies of 56.8 ± 29.0 and 39.1 ± 24.4 respectively. GPT-4 was non-inferior to the second human annotator in all datasets except kidney pathology. Small, open-source LLMs offer an accessible solution for the curation of local research databases; they obtain human-level accuracy while only leveraging desktop-grade hardware and ≤ 100 training reports. Unlike commercial LLMs, these tools can be locally hosted and version-controlled. <u>Strata</u> enables automated human-level performance in extracting structured data from clinical notes using ≤ 100 training reports and a single desktop-grade GPU.

## Introduction

Artificial intelligence (AI) tools in medicine aim to improve patient care by learning patterns across massive, well-annotated datasets. These annotations, describing key patient characteristics, are often derived from clinical notes, such as pathology and radiology reports. Extracting ground-truth labels from clinical notes remains a critical research bottleneck. The traditional approach to label curation is manual chart review, where researchers annotate thousands of reports. If the research team wishes to add a new variable to their database, a new round of exhaustive annotation is required. It is intractable to create health-system scale datasets with retrospective manual annotation. There is an unmet need for scalable and human-level data extraction from clinical reports.

Automated tools, including deep learning (1,2), statistical machine learning (3–6), and rule-based (7–9) methods, have been proposed to extract structured information from clinical reports. While these systems have achieved promising performance, they require substantial effort to develop and deploy. As a result, their adoption by the research community remains limited. Large language models (LLMs) have demonstrated exciting capabilities in general text understanding, detecting errors in radiology reports (10), streamlining radiology report impressions (11,12), and automatic synoptic reports (13). Despite the promising capabilities of commercial LLMs, they are ill-suited for large-scale research database curation due to reproducibility, cost, and privacy concerns. Commercial LLMs are continually updated.It is infeasible to reproduce prior extractions after model updates, and new versions of the LLM may unexpectedly perform worse when given the same inputs. Furthermore, costs associated with using commercial LLM can pose significant barriers, and institutional agreements are often required to address privacy concerns. Ideally, researchers could locally host their LLMs (“self-hosting”) on cheap “desktop-grade” hardware; these local LLMs would enable reproducible research while minimizing costs and avoiding the transfer of sensitive data.

Open-source LLMs, such as Llama (14–16) and Mistral (17), offer a promising solution for efficient “self-hosted” information extraction. They offer small LLM variants (i.e., <11 billion parameters) with instruction tuning, allowing for efficient general text understanding and cheap customization. Recent medically fine-tuned LLMs, including PMC-Llama 13B and Lama-3-8B-UltraMedical, have demonstrated compelling performance on biomedical questions answering benchmarks (18,19). Small versions of the DeepSeek-R1 model have demonstrated strong general reasoning performance while building on small-scale general LLM backbones (20), Llama-8B and Qwen-2.5-7B. While these LLMs offer exciting performance across general benchmarks, it remains unclear how these tools perform in clinical information extraction.

In this study, we focus on empowering individual researchers to curate large-scale research databases at their institution, using minimal computing (i.e., a few hours on a single GPU) and annotation resources (i.e., <100 reports annotated). Our strategy aligns with the practical constraints and foundational research workflows of academic medical centers. Following this strategy, we do not develop massive 400B+ open-source LLMs that require multiple H100 servers to host or generic universal LLMs that can generalize across institutions. We hypothesized that with simple fine-tuning techniques, small-scale open-source LLMs could enable individual researchers to create high-quality datasets customized to their institution, empowering their downstream AI research. Our extraction quality goal is “human-level performance”; specifically, we test the ability of LLMs to achieve statistically non-inferior performance to a research assistant extracting the same information from clinical notes. We benchmark a wide range of “self-hosted” LLM approaches on four diverse real-world datasets from ongoing AI research projects at our institutions. To facilitate the broader adoption of customized open-source LLMs, we release a low-code library, Strata, that supports the fine-tuning, evaluation, and deployment of LLMs for clinical information extraction.

## Materials and Methods

The local institutional review board approved this Health Insurance Portability and Accountability Act-compliant study and waived the requirement for written informed consent due to minimal risk and retrospective nature of the study with no subject contact. All procedures were carried out in accordance with the relevant guidelines and regulations of our university ethics committee and the Declaration of Helsinki.

We collected four diverse clinical text datasets underlying active research projects at our institutions across breast, kidney, prostate, and bone marrow cancer (Myelodysplastic Syndrome, MDS). These real-world datasets represent our intended use cases, where LLM-based tools empower the automatic creation of large-scale structured datasets to support downstream research at a single medical center. Our breast, kidney, and prostate datasets were annotated by a research assistant, and our MDS dataset was annotated by a hematologist. We describe each dataset in detail, including its creation and annotation process, in our supplementary methods.

### Extracting Structured Information using Customized Large Language Models

State-of-the-art LLMs are trained to follow instructions specified in text prompts when generating their answers. This process, called instruction-tuning (21), enables users to define tasks with instruction prompts. To leverage instruction-tuned LLMs for clinical information extraction, a user can write out the desired task (e.g., extract the cancer subtype from a pathology report) in a text prompt before inputting the full clinical report. Given the prompt and the clinical report, the LLM will generate an answer in the text format, which can be parsed into the desired structured format using a simple post-processing script. This process is illustrated in Figure 2.

**Figure 1.**
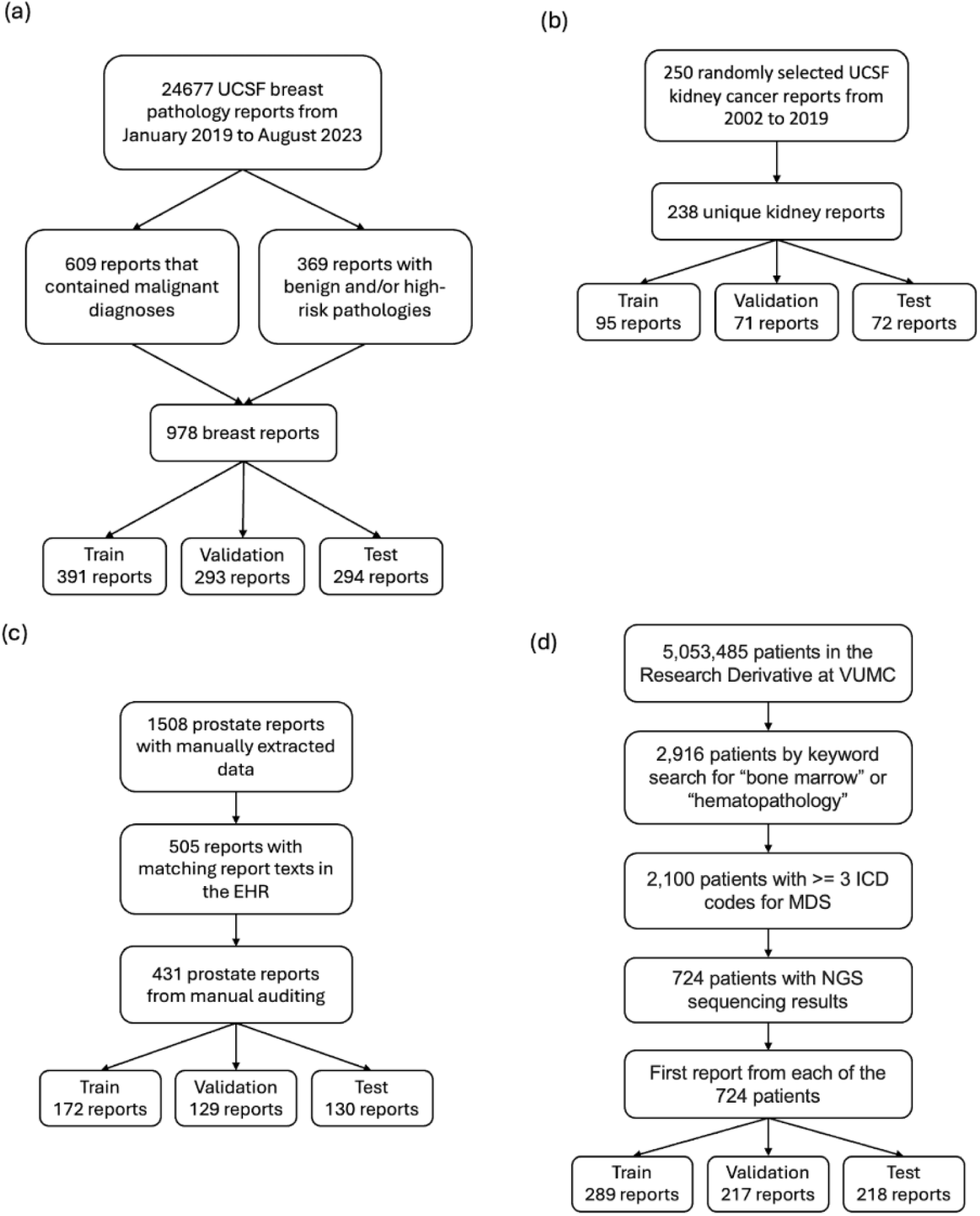
Dataset construction flowchart for our breast(a), kidney(b), prostate(c), and MDS (d) datasets. UCSF = University of California, San Francisco, VUMC = Vanderbilt University Medical Center, EHR = electronic health records, MDS = Myelodysplastic Syndrome.

**Figure 2.**
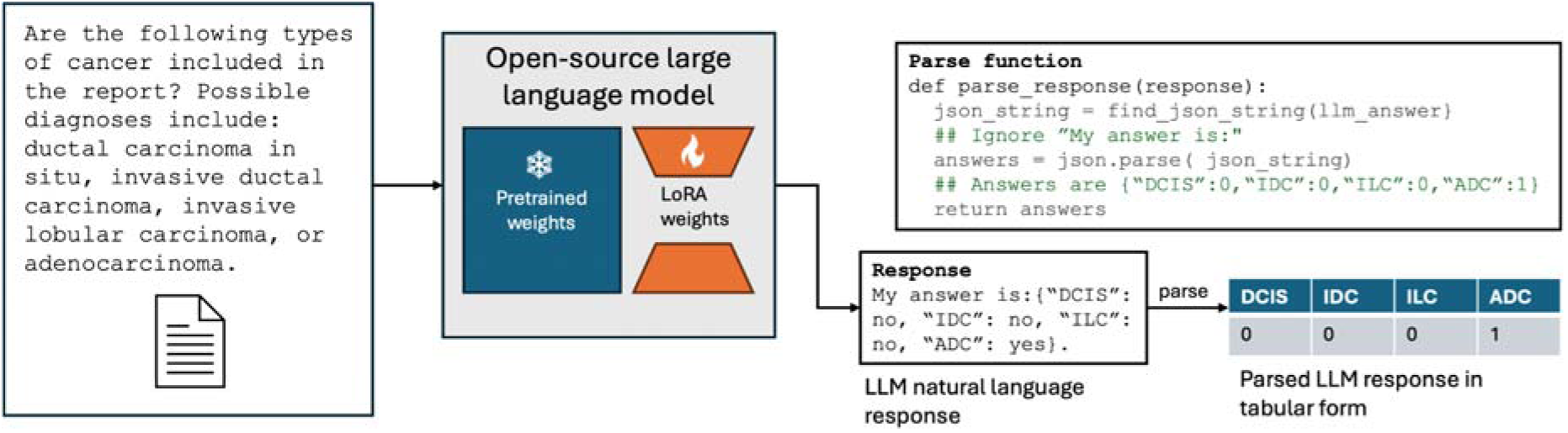
Process for leveraging open-source LLMs to extract structured information from clinical reports. The input to the LLM consists of a user prompt, which specifies the task (e.g., cancer subtyping), paired with the report text. An open-source LLM contains pre-trained weights (e.g., 8B parameters for Lamma-3.1 8B) learned across web-scale datasets; when fine-tuning an LLM, we introduce additional LoRA weights (∼400M parameters) to customize the model. The model generates an answer in text, parsed by a user-defined “parse function” into a tabular form.

In this work, we are interested in customizing local LLMs for clinical information extraction. To enable the full research community to benefit from this resource, we focus on LLMs that can be easily hosted on a single consumer desktop-grade GPU, such as the Nvidia RTX 3090 or 4090. As a result, we leverage three classes of small LLMs: 1) medically fine-tuned QA models, including PMC-LLaMA 13B and Llama-3-8B-UltraMedical, 2) accessible versions of leading reasoning models, including DeepSeek R1 distilled to Llama-3.1 8B and Qwen2.5-Math 7B, and 3) popular base instruction-tuned models, such as Llama-3.1 8B, Gemma-2-9B, and Mistral-v0.3 7B. These billion-parameter LLMs are orders of magnitude smaller than state-of-the-art LLMs such as Llama-3.1 405B and GPT-4. To enable smaller LLMs to achieve human-level performance in clinical information extraction, we fine-tune them. Specifically, we leverage simple preprocessing scripts to convert human-annotated structured labels (i.e., in a tabular format) to text answers for the LLM to generate (e.g., “My answer is: {“DCIS”: “yes”, “ILC”: “no”}”). Given a dataset of pairs of clinical reports and answer texts, we can train the LLM to correctly predict the intended answer from the user prompt and clinical report. This process is illustrated in Figure 3. To fine-tune LLMs using desktop-grade hardware, we employ Low-Rank Adaption (LoRA) (22). Instead of directly optimizing the billions of parameters in our LLMs, LoRA keeps the initial weights of the LLM frozen and adds a small number of new parameters (e.g., <400M parameters) across the LLM to customize the model’s behavior.

**Figure 3.**
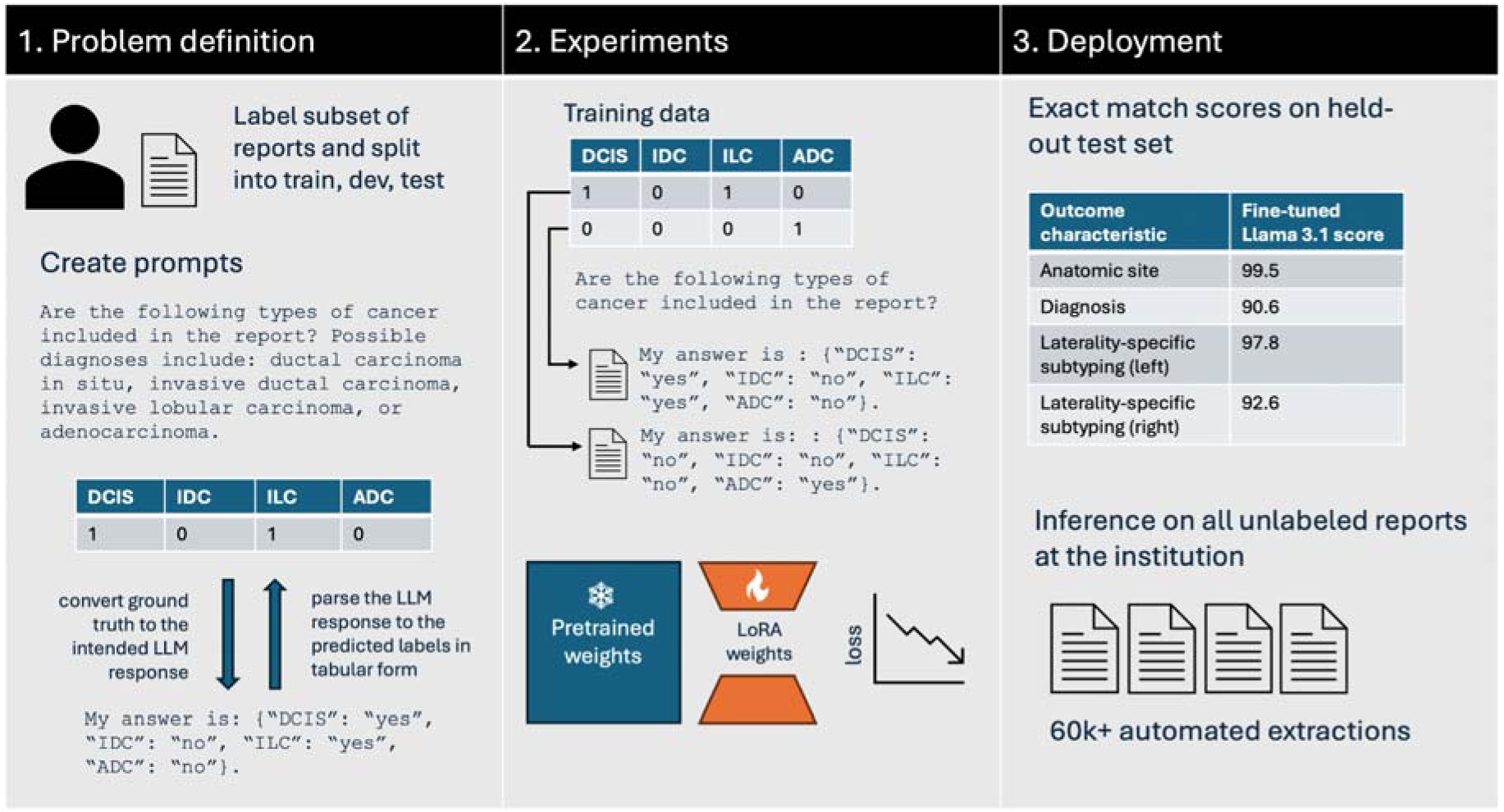
Process of using Strata to create research databases. The researcher labels a subset of the reports, defines prompts, and creates parsing functions to interpret the model’s response. Given this information, Strata supports LLM fine-tuning and deployment. In LoRA fine-tuning, the model’s LoRA weights are trained to generate the templated answers. Strata supports model evaluation and efficient bulk-inference. Bulk-inference is used to create a research database from all unlabeled reports at the institution.

To enable the broader clinical and research communities to fine-tune and deploy their LLMs on local hardware, we developed <u>Strata</u>, a lightweight library for clinical information extraction. As illustrated in Figure 3, Strata is used in three phases. First, researchers annotate a small dataset of clinical reports with their desired structured labels, which are partitioned into a training, development, and testing set. In addition to these annotations, researchers define the task for the LLM by specifying a prompt, a parsing script, and a template function. As described above, the parsing script maps the LLM’s answer to the desired structured format, and the template function maps the structured annotation to a text-based answer. Given an annotated dataset, Strata supports LoRA-based fine-tuning and flexible hyperparameter optimization. Finally, Strata offers a rich automated evaluation suite for model selection. We leverage Strata across all our experiments.

### LLM Development and Evaluation

Across all datasets, we developed prompts, parsing scripts, and template functions to maximize the performance of our open-source LLMs on the development sets. In these experiments, the models were used without fine-tuning (i.e. zero-shot). For the DeepSeek-R1 models, we modified the prompts to allow for additional reasoning preceding the final answer. Given a final set of prompts and scripts, we used LoRA to fine-tune our best-performing non-reasoning model, Llama-3.1.

LLM fine-tuning requires selecting multiple hyperparameters, which leads to a variety of design choices on model architecture, data augmentation, and training recipes. LoRA includes two hyperparameters, namely rank, and alpha. LoRA rank determines the size of the trainable parameters in fine-tuning, and alpha scales the output of these trainable layers relative to the original model’s outputs. In the context of the MDS dataset, we also experimented with class balancing, where rare values are oversampled during training. Finally, we also tuned the number of training epochs and the learning rate of the optimizer. Using Strata, we performed a grid search over possible hyperparameters in mixed-dataset fine-tuning on the development set. We applied the optimal hyperparameters in all other settings.

We considered three possible variants of LLM fine-tuning. First, we trained a customized LLM for each task within each dataset separately (i.e., question-specific fine-tuning). Next, we explored dataset-specific fine-tuning, in which all tasks within a single dataset (e.g., specimen site, cancer presence, and subtyping) were included in fine-tuning a model. Finally, we also considered training a single fine-tuned LLM across all datasets and tasks (i.e., mixed-dataset fine-tuning).

We leverage the annotations of our first human annotator as our “ground truth” in all evaluations. Our primary evaluation metric is “exact-match accuracy,” where we evaluate the ability of the LLM to extract all components of user-specified queries correctly. For instance, if the ground truth (i.e., first human annotator) response for breast cancer subtyping were “{DCIS: Yes, ILC: No, IDC: Yes},” the LLM predictions would have to match across all fields to gain a point in our cancer subtyping exact-match accuracy calculation. For dataset-level exact match accuracies, the LLM must correctly extract the information across all user queries for that clinical report. In addition to our stringent exact match accuracies, we also evaluate macro-F1, precision, and recall scores in our supplementary analyses.

We compare our open-source LLM results to GPT-4 and a second human annotator. All LLMs and the second human annotator were given the same prompts, and their extractions were compared to the first human annotator extractions as ground truth, using the metrics described above. The accuracy of our second human annotator serves as our benchmark for “human-level performance”. We hypothesize that fine-tuned LLMs would be non-inferior to the second human annotator in clinical information extraction. Our goal is not to outperform the second human annotator in matching the first human annotator; we expect disagreements between human annotators to reflect ambiguity in clinical reports. The second human annotator for each test set was a research assistant who received training on the relevant pathology classifications and the specific criteria for annotation for each dataset.

To understand the dependence of fine-tuned LLM performance on labeled data, we performed learning curve analyses. We randomly sampled subsets of each training set (consisting of 25% and 50% of the full training data) and fine-tuned using only the subsets at the dataset-specific level. We then evaluated the model performance with different subsets against the second human annotator to determine the number of labeled reports needed for human-level performance. To understand the effect of hyper-parameters on our performance, we performed a sensitivity analysis. For each hyperparameter (i.e., LoRA rank, LoRA scaling, number of epochs, and learning rate), we varied one hyperparameter value while holding the others constant. We evaluated the difference in exact match accuracy.

### Statistical Analysis

We used scikit-learn (version 1.5.1; scikit-learn.org) and SciPy (version 1.14.1; scipy.org) to compute all evaluation metrics, including F1-score, precision, and recall. We computed evaluation metrics across 5000 bootstrap samples (16) to obtain 95% confidence intervals (CIs). To compare the exact match accuracy of LLMs to the accuracy of a second human annotator, we utilized a non-inferiority t-test using the stats R package (version 4.4.1; cran.r-project.org). Given the size of each test set, 80% desired power, a 0.05 p-value, and the standard deviation of the second human annotator accuracy, we computed the margin of error measurable by each test set. This power calculation was performed using the pwr R package (version 1.3-0; *cran.r-* project.org), and we utilized dataset-specific margins in each non-inferiority calculation. Given 294, 130, 72, and 218 reports in the breast, prostate, kidney, and MDS test sets, this resulted in non-inferiority margins of 6.3%, 8.2%, 13.5%, and 5.9%, respectively.

## Results

### Report Characteristics

Our breast, kidney, prostate, and MDS datasets contained 978, 238, 431, and 724 reports (Figure 1). Of these reports, 294, 72, 130, and 218 were randomly selected for held-out testing for breast, kidney, prostate, and MDS extractions, and these reports contained an average of 766, 688, 423, and 785 words, respectively. Amongst breast, kidney, and prostate test sets, 65%, 94%, and 85% respectively contained a confirmation of cancer. 7% of the MDS test set confirmed untreated MDS. We report detailed text and label statistics for all four datasets in Table 1.

**Table 1.** Detailed Outcome and Text Characteristics for each Dataset.

| Outcome Characteristic | Train | Val | Test |
| --- | --- | --- | --- |
| **Breast** | n=391 | n=293 | n=294 |
| **No. of words** mean±SD (min-max) | 809±677 (200-3596) | 733±579 (189-3991) | 766 ± 640 (183-3717) |
| **Anatomic Site** |  |  |  |
| Right breast | 222 (57%) | 162 (55%) | 166 (56%) |
| Left breast | 213 (54%) | 158 (54%) | 150 (51%) |
| Right lymph node | 41 (10%) | 30 (10%) | 336 (114%) |
| Left lymph node | 41 (10%) | 29 (10%) | 34 (12%) |
| **Diagnosis** |  |  |  |
| Cancer | 242 (62%) | 177 (60%) | 190 (65%) |
| High risk | 31 (8%) | 29 (10%) | 23 (8%) |
| Benign | 118 (30%) | 87 (30%) | 81 (28%) |
| **Laterality-specific subtyping** |  |  |  |
| Ductal carcinoma in situ | 142 (36%) | 109 (37%) | 121 (41%) |
| Invasive ductal carcinoma | 109 (28%) | 91 (31%) | 109 (37%) |
| Invasive lobular carcinoma | 25 (6%) | 16 (5%) | 12 (4%) |
| Adenocarcinoma NOS | 12 (3%) | 12 (4%) | 5 (2%) |
| **Kidney** | n=95 | n=71 | n=72 |
| **No. of words** mean±SD (min-max) | 768±412 (270-3081) | 741±361 (275-2081) | 688±259 (128-1652) |
| **Cancer Diagnosis** |  |  |  |
| Yes | 84 (88%) | 62 (87%) | 68 (94%) |
| No | 11 (12%) | 9 (13%) | 4 (6%) |
| **Cancer Anatomic Site** |  |  |  |
| Left lower pole | 19 (20%) | 12 (17%) | 11 (15%) |
| Right lower pole | 10 (11%) | 8 (11%) | 11 (15%) |
| Left middle pole | 11 (12%) | 14 (20%) | 11 (15%) |
| Right middle pole | 17 (18%) | 10 (14%) | 15 (21%) |
| Left upper pole | 17 (18%) | 17 (24%) | 16 (22%) |
| Right upper pole | 13 (14%) | 10 (14%) | 13 (18%) |
| **Prostate** | n=172 | n=129 | n=130 |
| **No. of words** mean±SD (min-max) | 431±131 (110-916) | 436±176 (166-1587) | 423±109 (172-699) |
| **PIRADS** |  |  |  |
| 5 | 58 (34%) | 32 (25%) | 43 (33%) |
| 4 | 69 (40%) | 54 (42%) | 46 (35%) |
| 3 | 15 (9%) | 18 (14%) | 20 (15%) |
| 2 | 14 (8%) | 12 (9%) | 12 (9%) |
| 1 | 3 (2%) | 2 (2%) | 0 (0%) |
| **Cancer Diagnosis** |  |  |  |
| Yes | 149 (87%) | 108 (84%) | 111 (85%) |
| No | 23 (13%) | 21 (16%) | 19 (15%) |
| **MDS** | n=289 | n=217 | n=218 |
| **No. of words** mean ± SD (min-max) | 810±222 (219-1652) | 805±234 (171-1725) | 785±225 (151-1612) |
| **Bone marrow** |  |  |  |
| Yes | 254 (88%) | 195 (90%) | 193 (89%) |
| No | 35 (12%) | 22 (10%) | 25 (11%) |
| **Untreated Myelodysplastic Syndrome** |  |  |  |
| Yes | 19 (7%) | 18 (8%) | 16 (7%) |
| No | 235 (81%) | 177 (82%) | 177 (81%) |
| Missing | 35 (12%) | 22 (10%) | 25 (12%) |

Note – Unless otherwise specified, data are numbers of reports, with percentages for each split in parentheses. For breast and prostate, “laterality-specific” means that the model was asked separately for each laterality (see Table 2). For diagnosis characteristics, the outcomes sum to 100%. For anatomic site and subtyping, the outcomes are not mutually exclusive, so the outcomes sum to >100%. NOS = not otherwise specified.

### LLM Performance

We report our prompts for each dataset and task in Table 2 and a detailed model performance evaluation in Table 3. Our primary performance metric, exact match accuracy, evaluates the ability of a model to correctly extract all fields for a report, using the labels of the first human annotator as the “ground truth.” We compare model accuracies to the accuracies of independent second human annotators using non-inferiority tests. We consider a model to have obtained “human-level performance” if it meets the accuracy of our second human annotator. Our second human annotators obtained exact match accuracies of 75.5 (95% CI 70.4, 80.3), 83.3 (95% CI 73.6, 90.3), 83.1 (95% CI 75.4, 88.5), 85.8 (95% CI 80.7, 89.9) in the breast, kidney, prostate, and MDS test set, respectively. These correspond to average inter-annotator agreement scores of 0.87, 0.78, 0.88, and 0.54 for each dataset. When leveraging dataset-specific fine-tuning, Llama-3.1 8B obtained exact-match accuracies of 91.5 (95% CI 88.1, 94.2), 87.5 (95% CI 77.8, 94.4), 91.5 (95% CI 85.4, 95.4), 89.4 (95% CI 84.9, 93.1) in the breast, kidney, prostate, and MDS test set, respectively. This model obtained higher accuracies than the second human annotator in all four datasets, and the result was statistically non-inferior in all test sets (p<0.001). These results are summarized in Figure 4, and we report additional detailed metrics by data field in Table 4. We report additional metrics, including precision, recall, and F1 scores, in the supplementary results.

**Figure 4.**
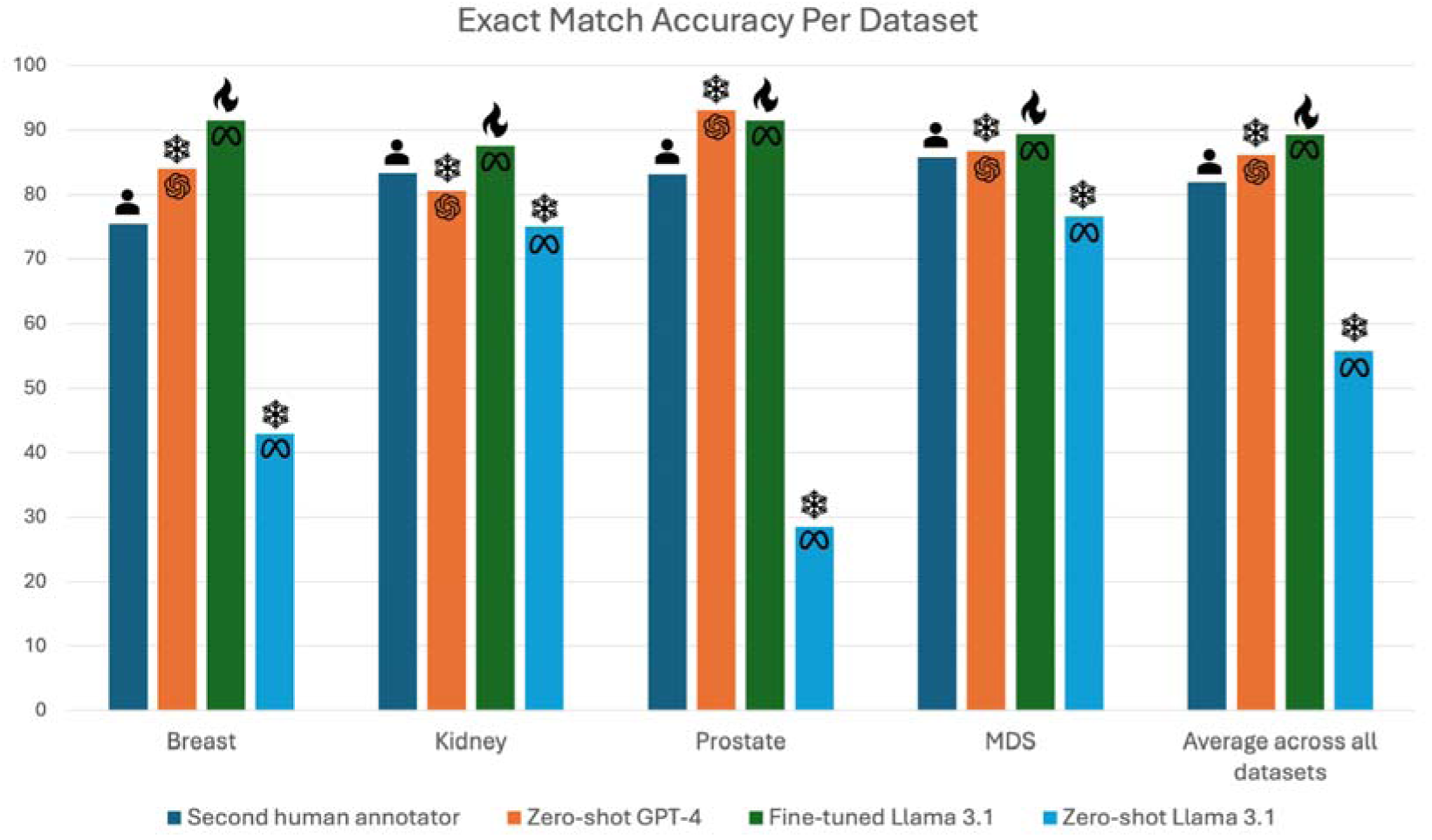
Summarized dataset-level results. Bar graph shows exact match accuracy for each dataset. Selected models include the second human annotator, zero-shot GPT-4, and fine-tuned and zero-shot Llama 3.1 8B. The human, snowflake, and flame icons represent the human, zero-shot, and fine-tuned models respectively.

**Table 2.**
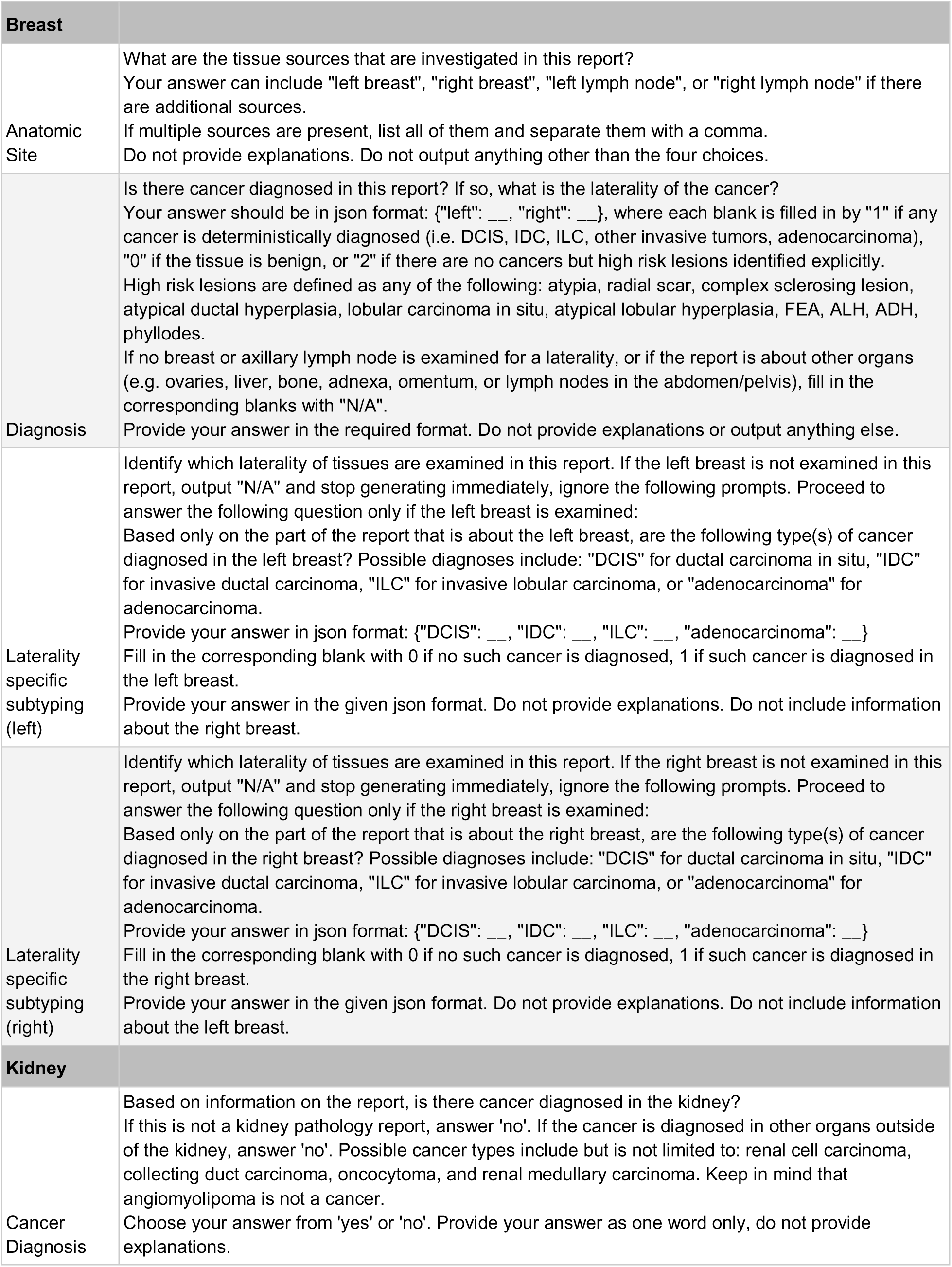
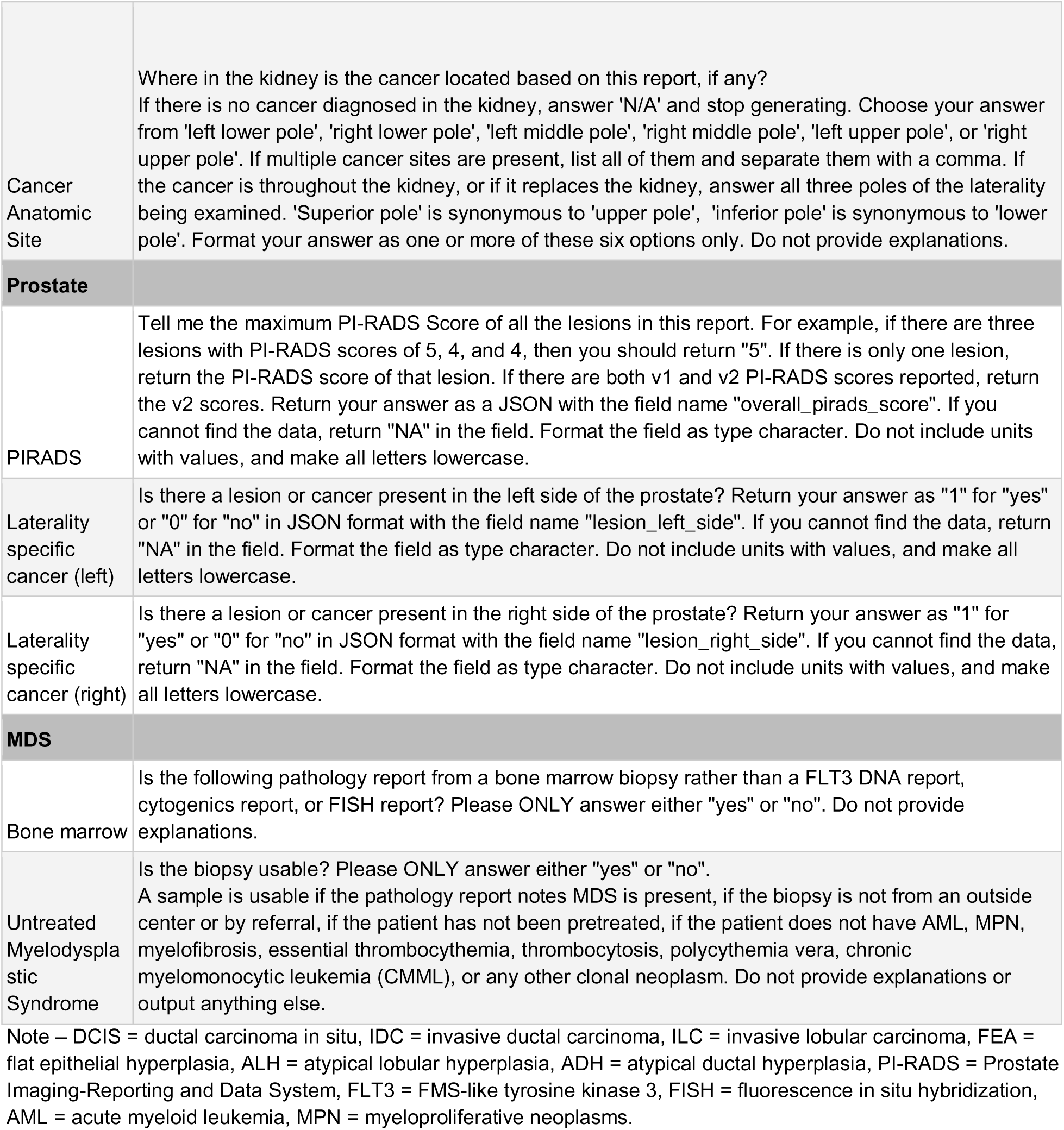
Task-specific LLM Prompts for each Dataset.

**Table 3.** Dataset-Level Exact Match Accuracies for the Second Human Annotator, GPT-4 and Open-source LLMs.

|  |  | Closed-source zero shot | Open-source zero-shot |  |  |  |  |  | Question-specific training | Dataset-specific training | Mixed-dataset training |
| --- | --- | --- | --- | --- | --- | --- | --- | --- | --- | --- | --- |
|  | Second Human Annotator | GPT-4 | Llama-3-8B-UltraM edical | Meta-Llama-3.1-8B-Instruct | Mistral-7B-Instruct-v0.3 | Gemma-2-9b-it | DeepSeek-r1-distill-llama-8b | DeepSeek-r1-distill-qwen-7b | Meta-Llama-3.1-8B-Instruct | Meta-Llama-3.1-8B-Instruct | Meta-Llama-3.1-8B-Instruct |
| Breast | 75.5<br>[70.4, 80.3] | 84.0<br>[79.6, 87.8] | 27.6<br>[22.4, 32.9] | 42.9<br>[79.5, 83.8] | 29.3<br>[26.5, 34.7] | 65.6<br>[60.2, 71.1] | 72.4<br>[67.3, 77.6] | 36.7<br>[31.3, 42.5] | 89.1<br>[85.4, 92.3] | **91.5**<br>**[87.8, 94.2]** | 90.8<br>[87.1, 93.9] |
| Kidney | 83.3<br>[73.6, 90.3] | 80.6<br>[70.8, 88.9] | 44.4<br>[33.3, 55.6] | 75.0<br>[63.9, 83.3] | 72.2<br>[61.1, 81.9] | 12.5<br>[8.8, 20.2] | 66.7<br>[55.6, 77.8] | 41.7<br>[30.6, 54.2] | 80.6<br>[70.8, 88.9] | **87.5**<br>**[77.8, 94.4]** | 73.6<br>[62.5, 81.9] |
| Prostate | 83.1<br>[75.4, 88.5] | **93.1**<br>**[87.7, 96.2]** | 75.4<br>[67.7, 82.3] | 28.5<br>[59.5, 69.2] | 76.9<br>[69.2, 83.8] | 84.6<br>[77.7, 90.0] | 80.8<br>[73.8, 86.9] | 53.8<br>[45.4, 62.3] | 90.8<br>[84.6, 94.6] | 91.5<br>[86.2, 95.4] | 90.8<br>[84.6, 95.4] |
| MDS | 85.8<br>[80.7, 89.9] | 86.7<br>[81.7, 90.8] | 9.2 [6.0, 13.8] | 76.6<br>[84.2, 90.6] | 16.1 [8.7, 17.4] | 45.9<br>[39.4, 52.8] | 7.3 [4.6, 11.5] | 20.6<br>[15.6, 26.6] | 86.7<br>[81.7, 90.8] | **89.4**<br>**[84.9, 93.1]** | 86.7<br>[81.7, 90.8] |
| Average | 81.9 ± 3.9 | 86.1 ± 4.6 | 39.1 ± 24.4 | 55.8 ± 20.7 | 48.6 ± 26.4 | 52.1 ± 26.7 | 56.8 ± 29.0 | 38.2 ± 11.9 | 86.8 ± 3.9 | **90.0 ± 1.7** | 85.5 ± 7.1 |

Note – Dataset-level exact match accuracies are followed by 95% confidence intervals in brackets. This metric evaluates the ability of each model to correctly extract all components from a report. We report the average exact match accuracy across all four datasets, in addition to standard deviation, in the “Average across all datasets” row.

**Table 4.** Detailed comparison of fine-tuned Llama 3.1 to Second Human Annotator.

|  | Fine-tuned Llama 3.1 8B | Second human annotator | Non-inferiority p-values |
| --- | --- | --- | --- |
| **Breast** |  |  |  |
| Anatomic Site | 99.0 [97.3, 99.7] | 87.8 [84.0, 91.2] |  |
| Diagnosis | 95.6 [92.9, 97.6] | 92.9 [89.5, 95.2] |  |
| Laterality-specific subtyping (left) | 99.3 [97.6, 100.0] | 94.2 [91.2, 96.3] |  |
| Laterality-specific subtyping (right) | 96.6 [93.9, 98.3] | 95.2 [92.5, 97.3] |  |
| Overall exact match | 91.5 [87.8, 94.2] | 75.5 [70.1, 80.3] | < 0.001 |
| **Kidney** |  |  |  |
| Cancer Diagnosis | 98.6 [93.1, 100.0] | 93.1 [86.1, 97.2] |  |
| Cancer Anatomic Site | 87.5 [77.8, 94.4] | 86.1 [76.4, 93.1] |  |
| Overall exact match | 87.5 [77.8, 94.4] | 83.3 [73.6, 90.3] | <0.001 |
| **Prostate** |  |  |  |
| PIRADS | 97.7 [93.8, 99.2] | 91.5 [85.4, 95.4] |  |
| Cancer (left) | 96.2 [91.5, 98.5] | 93.1 [87.7, 96.9] |  |
| Cancer (right) | 96.2 [91.5, 98.5] | 95.4 [90.8, 98.5] |  |
| Overall exact match | 91.5 [85.4, 95.4] | 83.1 [76.2, 88.5] | < 0.001 |
| **MDS** |  |  |  |
| Bone marrow | 91.7 [87.6, 95.0] | 92.7 [88.5, 95.9] |  |
| Untreated Myelodysplastic Syndrome | 96.9 [93.3, 99.0] | 92.2 [87.6, 95.3] |  |
| Overall exact match | 89.4 [84.9, 93.1] | 85.8 [80.3, 89.9] | < 0.001 |

Note – Data are exact match accuracies with 95% confidence intervals in brackets. Overall exact match accuracy measures if exactions across all tasks for a particular report were correct. The p-values are calculated with non-inferiority margins based on the size of each test set, 80% power, and a significance threshold of 0.05. This resulted in non-inferiority margins of 6.3%, 13.5%, 8.2%, and 5.9% for breast, kidney, prostate, and MDS, respectively.

Without fine-tuning (i.e. zero-shot), open-source LLMs obtain significantly worse performance than our second human annotator, who had an average exact match accuracy of 81.9%. DeepSeek-R1-Distill-Llama-8B obtains the highest overall performance, with an average exact match accuracy of 56.8% and 72.4 (95% CI 67.3, 77.6), 66.7 (95% CI 55.6, 77.8), 80.8 (95% CI 73.8, 86.9), and 7.3 (95% CI 4.6, 11.5) on breast, kidney, prostate and MDS, respectively. Llama-3.1 8B obtained similar zero-shot performance averaging 55.8 over the four datasets, and 42.9 (95% CI 79.5, 83.8), 75.0 (95% CI 63.9, 83.3), 28.5 (95% CI 59.5, 69.2), and 76.6 (95% CI 84.2, 90.6) for each dataset. Critically, Llama-3.1 8B obtained this performance without the use of any intermediate reasoning steps and, hence, was readily amenable to further fine-tuning. Gemma-2-9B and Mistral-v0.3 7B both obtained worse performance than Lamma-3.1 8B, with average exact match accuracies of 52.1% and 48.6%, respectively. We found that LLama-3-8B-UltraMedical performed worse than Llama 3.1 8B, with 39.1 average exact match across datasets. We do not report numbers for PMC-LLaMA 13B because the model failed to generate properly formatted outputs and our clinical notes exceeded the model’s maximum sequence length. Llama-3.1 8B obtained qualitatively similar results across fine-tuning settings, obtaining average exact match accuracies ranging from 85.5% to 89.3% compared to 81.9% by second human annotators. These results suggest that researchers can obtain human-level results if they annotate data for a single task (i.e., question-specific finetuning); alternatively, institutions can train a single model to support all tasks (i.e., mixed-dataset finetuning) to obtain human-level results. Dataset-specific fine-tuned Llama-3.1 8B significantly outperformed its zero-shot counterparts across all datasets (p<0.05). GPT-4, which received no fine-tuning, obtained an average exact match accuracy of 86.1% across the four datasets. GPT-4 was non-inferior (p<0.001) to the second human reader in breast, prostate, and MDS test sets. GPT-4 did not obtain non-inferior accuracy relative to the second human annotator (80.6% vs 83.3%, p=0.09) in the kidney test set.

### Learning Curve Analysis and Hyperparameter Sensitivity

To understand the minimum number of training samples required to achieve human-level performance, we retrained our dataset-specific fine-tuned Llama-3.1 8B model with subsets of our training set (Figure 5). Across each test set, we report the performance of our second human annotator on our dotted line. Our fine-tuned Llama-3.1 8B matched human performance when leveraging only 100, 95, 43, and 72 training reports for the breast, kidney, prostate, and MDS test sets, respectively. Each dataset-level model took 1-3 hours to train using a single A40 GPU. At the time of writing, these experiments would cost between $0.80 to $2.40 on a private cloud. Next, we assessed the sensitivity of our models to the optimal choice of training hyper-parameters, including learning rate, number of training epochs, LoRa scaling (i.e., alpha), and LoRa rank (Figure 6). We found that our fine-tuning results are sensitive to the choice of learning rate; while leveraging a learning rate of 1e-4 obtains a dataset-average exact-match accuracy of 89.3%, learning rates of 1e-3 and 1e-5 obtain degraded accuracies of 5.0% and 80.3%, respectively.

**Figure 5.**
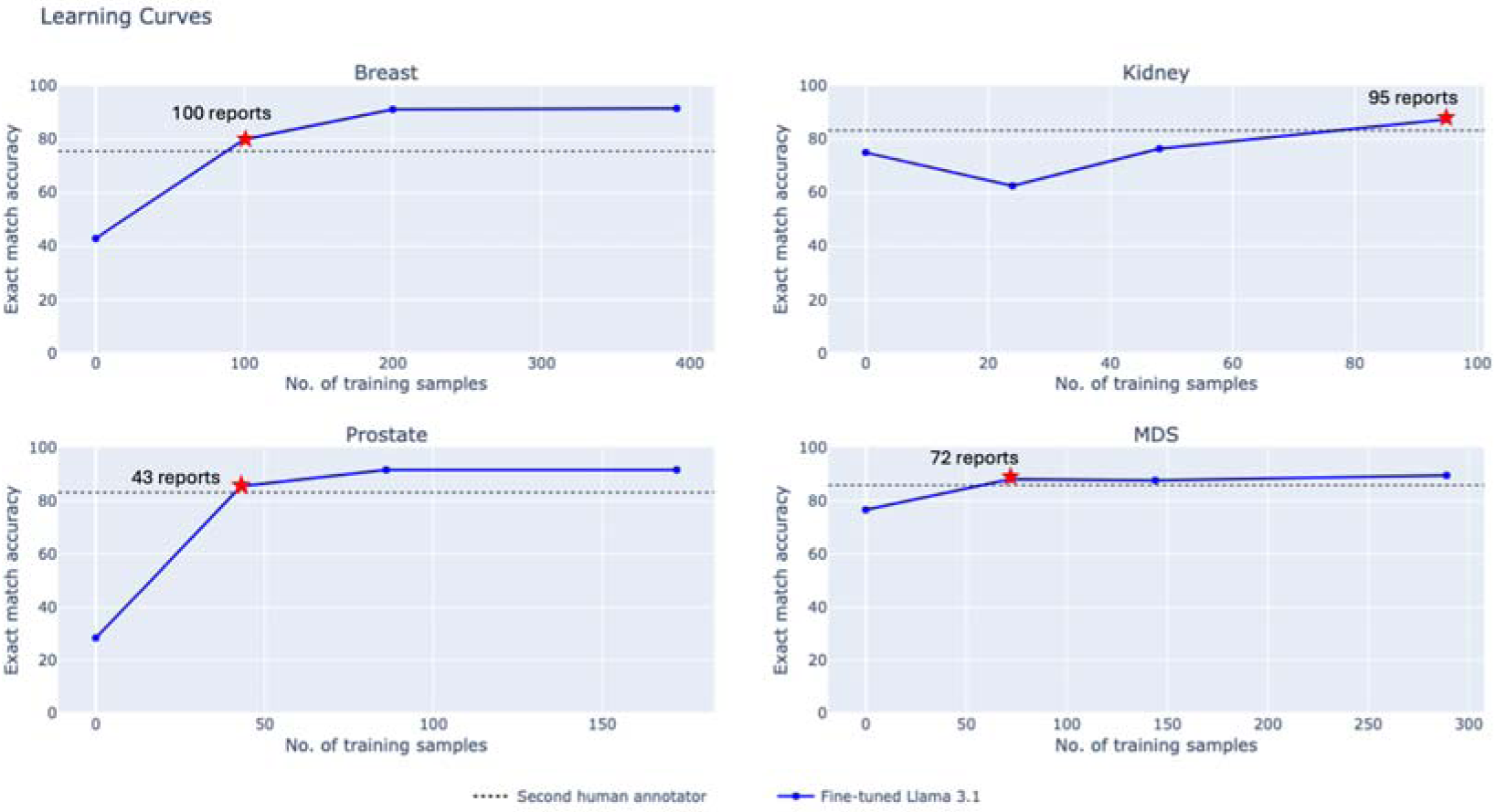
Learning curves analysis across all four datasets. Blue line graphs show the exact match accuracy for dataset-specific finetuned Llama 3.1 when leveraging 0%, 25%, 50%, and 100% of the training data. The dotted line displays the performance of the second human annotator. The stars show the number of reports used to match performance of the second human annotator.

**Figure 6.**
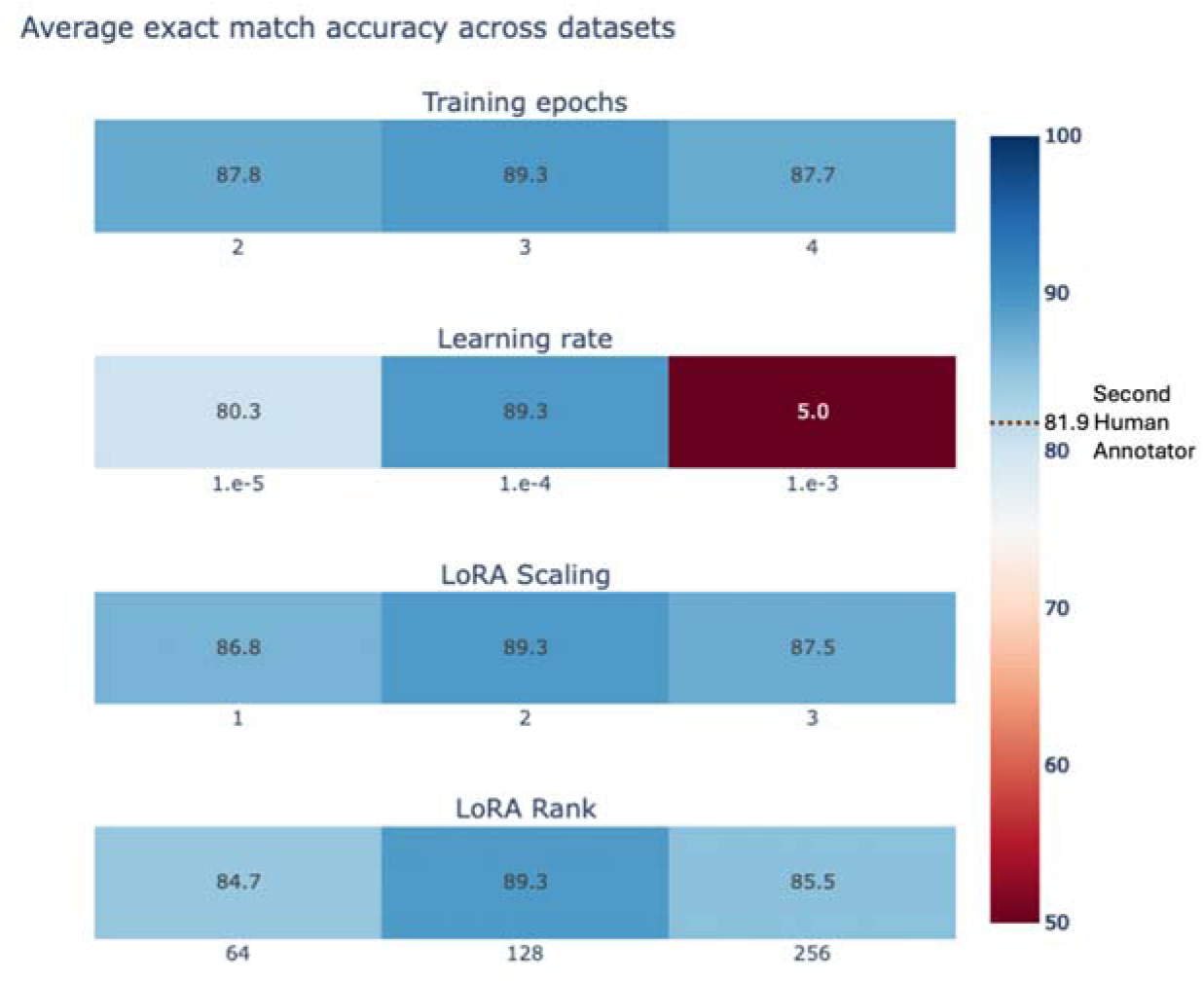
Hyperparameter sensitivity analysis for mixed-dataset fine-tuned Llama 3.1 8B. Heat maps display the average dataset-level exact match accuracy across datasets, across a range of values for each hyper-parameter. Llama 3.1 is robust to the choice of training epochs, LoRA (low-rank adaptation) scaling, and LoRA rank, but the choice of learning rate can significantly alter performance.

## Discussion

We developed Strata, a lightweight open-source library for LLM-based clinical information extraction. Strata supports a wide range of LLMs, prompt development, parameter-efficient fine-tuning, model evaluation, and deployment. Using Strata and minimal compute resources, namely a single commodity GPU (Nvidia A40) per experiment, we demonstrated that fine-tuned open-source LLMs achieve human-level performance in diverse, real-world clinical research applications across prostate MRI, breast pathology, kidney pathology, and myelodysplastic syndrome pathology reports. Our fine-tuned Llama-3.1 8B model was non-inferior to a second human annotator in all test sets. Moreover, Llama-3.1 8B demonstrated human performance leveraging only 100, 95, 43, and 72 training reports in our breast, kidney, prostate, and MDS test sets. These results demonstrate that locally hosted fine-tuned LLMs offer a practical solution for clinical information extraction, requiring limited computational and human effort; our “low-code” open-source library is designed to empower individual researchers to curate high-quality (i.e., human-level) databases at their instruction, supporting their downstream AI research.

Much of the prior work on using LLMs for clinical information extraction has focused on commercial models such as ChatGPT (23). In our study, we found that GPT-4 demonstrated human-level zero-shot performance in three out of four test sets, namely breast, prostate, and MDS, and it was not non-inferior to our second annotator in the kidney dataset. Relying on commercial models to develop research databases poses several challenges. Using commercial models for protected health information requires institutional agreements to address privacy concerns, creating a significant access barrier. Moreover, rigorous research practices require reproducibility; as commercial LLMs are updated without user oversight, researchers cannot easily reproduce prior model queries or ensure consistent extraction quality. In contrast, our small open-source LLMs can be locally hosted, version-controlled, and easily customized for new applications.

While there has been rapid progress in adapting open-source LLMs to medical (18,19) and general reasoning tasks (20), we found that both biomedically fine-tuned LLMs and general reasoning models did not outperform the base instruction-tuned Llama-3.1 8B model in clinical information extraction. Specifically, we found that Llama-3-8B-UltraMedical obtained substantially worse performance than Llama-3.1 8B, and PMC-Llama 13B failed to general well-formatted outputs. Leading reasoning LLMs, including deepseek-r1-distill-llama-8b, obtained similar performance to Llama-3.1 8B, despite the use of intermediate reasoning steps. In contrast, our fine-tuned Llama-3.1 significantly exceeded the zero-shot Llama-3.1 model, obtaining human-level performance.

We found that open-source LLMs address the limitations of traditional NLP techniques. Trivedi et al. (24) previously evaluated the performance of conventional NLP techniques applied to breast pathology reports and found that performance was lower for cases over 1024 characters with an F-measure of 0.83. In contrast, we saw consistent performance across our datasets, where all reports across all test sets contained over 1024 characters. Yala et al. (6) developed statistical machine-learning tools for parsing breast pathology reports; while the tool achieved an exact-match accuracy of 90%, it required over 6000 annotations to reach this performance. In contrast, we demonstrated that we could achieve human-level results leveraging only ≤100 training set reports, addressing the primary limitation of this prior work.

Our study has limitations. We used datasets from two tertiary academic institutions, which may not account for variability in pathology report structures, terminology, and style across different institutions. Our clinical information extraction tasks only capture a fraction of the data elements in clinical reports. Additional work is needed to broaden our results to more tasks and institutions.

## Conclusion

Our study shows that fine-tuned open-source LLMs can achieve human-level performance while leveraging modest annotation (i.e., ≤100 reports) and computational resources (i.e., <$3.00). Self-hosted open-source LLMs offer a compelling and high-performance alternative to commercial LLMs. We release Strata, our lightweight library, to broaden access to these tools.

## Data Availability

The institutional data used in this study are not publicly available due to compliance with patient privacy protection but are available from the corresponding author on reasonable request.

## Code Availability

The underlying code for this study is available at https://github.com/YalaLab/strata.

## Data Availability

All data produced in the present work are contained in the manuscript.

## Acknowledgements

We want to thank Trevor Darrell for thoughtful feedback on this study and Li-Ching Chen and Natalia Harguindeguy for beta-testing Strata. We also want to thank the UCSF Facility of Advanced Computing team, including Hunter McCallum, Sandeep Giri, Rhett Hillary, Marissa Jules, Sean Locke, and John Gallias, for their invaluable work in supporting our computational environment.

