## Appendix for "Human-level information extraction from clinical reports with fine-tuned language models" (from another version of this preprint: version 1, 2024-11-20)

**Supplementary Methods: Detailed Dataset Construction**

*Breast Pathology Dataset*
We performed a single-institution retrospective analysis of breast pathology reports from January 2019 to August 2023 using mPower Clinical Analytics. A total of 978 pathology reports were selected, including 609 reports that contained malignant diagnoses and 369 reports with benign and/or high-risk pathologies, to form a cancer-enriched dataset. High-risk pathologies included atypical intraductal epithelial proliferations (atypical ductal hyperplasia), lobular neoplasias (lobular carcinoma in situ and atypical lobular hyperplasia), flat epithelial atypia, radial scar/complex sclerosing lesion, and phyllodes tumor.

For each report, the readers annotated the sample source (right breast, left breast, right lymph node, and/or left lymph node) and diagnosis category (benign, high-risk, or cancer). If more than one pathology diagnosis was present from a sample source, the highest level (malignant > high risk > benign) was annotated. For malignant pathologies, cancer subtypes were further categorized as ductal carcinoma in situ, invasive ductal carcinoma, invasive lobular carcinoma, adenocarcinoma not otherwise specified, or other invasive malignant pathology. The dataset was split into 489/244/245 for training, validation, and testing.

*Kidney Pathology Dataset*

We performed a single-institution retrospective analysis of 250 randomly sampled kidney pathology reports from 2002 to 2019. For each report, the readers annotated the presence and site of renal malignancy. Malignant renal tumors included renal cell carcinoma, collecting duct carcinoma, oncocytoma, and renal medullary carcinoma. For malignant pathologies, the specific location of the cancer within the kidney was annotated as right upper pole, right middle pole, right lower pole, left upper pole, left middle pole, and/or left lower pole. If the site of malignancy was not specified, the report was annotated as “not specified.” Additionally, if the cancer was reported to involve the ureter, renal sinus, renal vein, veins, hilum, lymphatics, adrenal gland, or capsule/perirenal fat, then “other” was annotated in addition to any site noted within the kidney. The dataset was split 95/71/72 for training, validation, and testing.

*Prostate MRI Dataset*

The [Institution 1] Urology Department maintains a database of manually annotated prostate cancer MRIs from which data from 1508 reports were pulled. We identified 505 matching report texts in the EHR which were pulled for LLM analysis. Of these, 431 were from after 2015, which was when MRI reporting format became more standardized at [Institution 1]. For each prostate MRI report, the readers annotated the presence of a lesion or cancer for both the right and left sides. The maximum PI-RADS score of all lesions in the report was recorded (e.g., "5" if the scores are 5, 4, and 4). In cases where both v1 and v2 PI-RADS scores were reported, the v2 scores were utilized. The dataset was split 172/129/130 for training, validation, and testing.

*Myelodysplastic Syndrome (MDS) Dataset*

We searched for the occurrence of the keywords “bone marrow” or “hematopathology” at least once in 5,053,485 patients in the Research Derivative database at [Institution 2] and created a cohort of 2,916 patients. From this cohort, we then only included 2,100 individuals whose chart included three or more occurrences of ICD10 codes for myelodysplastic syndrome (D46 and its components). We then excluded individuals who did not have next-generation sequencing results to create the final cohort of 724 patients. We then selected the first pathology report from these 724 patients to increase the likelihood that reports described myelodysplastic syndrome before treatment or transformation to acute myeloid leukemia. Our final dataset included 724 unique reports.

Each report was then manually reviewed by a hematologist to indicate if they were associated with a bone marrow biopsy and if they were usable. Our annotator indicated whether the specimen was from a bone marrow biopsy. Next, our annotator determined if the report described untreated MDS and was relevant for a downstream research question. Specifically, a report was considered as containing “untreated MDS” if the pathology report indicated the presence of myelodysplastic syndrome, if the biopsy was not obtained from an outside center or by referral, and if the patient had not been pretreated or diagnosed with acute myeloid leukemia, myeloproliferative neoplasm, myelofibrosis, essential thrombocythemia, thrombocytosis, polycythemia vera, chronic myelomonocytic leukemia, or any other clonal neoplasm.

*Dataset Labeling*
In addition to the original annotations described above, clinical reports in each test set received an additional annotation by an independent research assistant (i.e., a second human annotator) who did not participate in the original annotation process. Research assistants received training on the relevant pathology classifications and the specific criteria for annotation for each dataset.

**Supplementary Methods: LLM fine-tuning hyperparameters**

LLM fine-tuning requires selecting multiple hyperparameters, which leads to a variety of design choices on model architecture, data augmentation, and training recipes. LoRA includes two hyperparameters, namely rank, and alpha. LoRA rank determines the size of the trainable parameters in fine-tuning, and alpha scales the output of these trainable layers relative to the original model's outputs. In the context of the MDS dataset, we also experimented with class balancing, where rare values are oversampled during training. Finally, we also tuned the number of training epochs and the learning rate of the optimizer. Using Strata, we performed a grid search over possible hyperparameters in mixed-dataset fine-tuning on the development set. We applied the optimal hyperparameters in all other settings.

**Supplementary Tables**

**Supplemental Table 1. Detailed evaluation metrics for second human annotator**

| *Question* | *Precision* | *Recall* | *F1* |
| --- | --- | --- | --- |
| **Breast** | | | |
| Anatomic Site | 99.8 [99.3, 100.0] | 77.6 [71.1, 83.0] | 86.1 [80.6, 90.1] |
| Diagnosis | 89.7 [83.4, 95.0] | 90.5 [83.9, 95.8] | 90.0 [84.6, 94.4] |
| Laterality specific cancer subtyping (left) | 84.4 [69.7, 97.2] | 91.4 [64.9, 97.5] | 85.6 [65.2, 95.1] |
| Laterality specific cancer subtyping (right) | 88.3 [66.4, 98.5] | 97.2 [71.2, 98.7] | 92.1 [68.8, 98.0] |
| Average | 90.6 ± 6.6 | 89.2 ± 8.3 | 88.4 ± 3.1 |
| **Kidney** | | | |
| Cancer Diagnosis | 98.5 [90.6, 100.0] | 94.1 [86.2, 98.5] | 96.2 [91.6, 98.6] |
| Cancer Anatomic Site | 81.0 [68.7, 86.5] | 84.4 [75.3, 91.3] | 81.7 [73.1, 87.1] |
| Average | 89.7 ± 12.4 | 89.3 ± 6.9 | 89 ± 10.3 |
| **Prostate** | | | |
| Laterality specific cancer diagnosis (left) | 92.1 [84.2, 97.2] | 95.9 [89.0, 98.7] | 94.0 [88.9, 97.2] |
| Laterality specific cancer diagnosis (right) | 96.1 [89.2, 98.8] | 96.1 [89.5, 98.8] | 96.1 [91.7, 98.6] |
| PI-RADS | 78.7 [76.4, 79.6] | 79.1 [77.1, 80.0] | 78.9 [77.0, 79.8] |
| Average | 89 ± 9.1 | 90.4 ± 9.8 | 89.7 ± 9.4 |
| **MDS** | | | |
| Bone marrow | 92.3 [88.0, 95.3] | 100.0 [nan, nan] | 96.0 [93.7, 97.6] |
| Untreated Myelodysplastic Syndrome | 52.0 [32.2, 72.7] | 81.2 [54.1, 95.2] | 63.4 [43.8, 78.8] |
| Average | 72.2 ± 28.5 | 90.6 ± 13.3 | 79.7 ± 23.1 |
| **Overall** | | | |
| Average | 85.4 ± 8.8 | 89.9 ± 0.7 | 86.7 ± 4.7 |

Dataset-level statistics include precision, recall, and F1 scores with 95% confidence intervals in brackets. Averages ± standard deviations per dataset are calculated across questions in each dataset, and overall averages ± standard deviations are calculated across the dataset averages.

**Supplemental Table 2. Detailed evaluation metrics for Llama 3.1 with dataset-specific fine-tuning**

| *Question* | *Precision* | *Recall* | *F1* |
| --- | --- | --- | --- |
| **Breast** | | | |
| Anatomic Site | 99.2 [96.7, 100.0] | 99.8 [99.3, 100.0] | 99.5 [98.4, 99.9] |
| Diagnosis | 94.8 [87.4, 98.8] | 87.6 [80.2, 94.0] | 90.6 [84.3, 95.6] |
| Laterality specific cancer subtyping (left) | 100.0 [nan, nan] | 95.9 [66.7, 100.0] | 97.8 [69.8, 100.0] |
| Laterality specific cancer subtyping (right) | 89.5 [67.8, 99.4] | 97.0 [72.1, 98.8] | 92.6 [70.1, 98.4] |
| Average | 95.9 ± 4.8 | 95.1 ± 5.2 | 95.1 ± 4.2 |
| **Kidney** | | | |
| Cancer Diagnosis | 97.1 [89.9, 100.0] | 100.0 [nan, nan] | 98.6 [94.6, 100.0] |
| Cancer Anatomic Site | 83.0 [71.7, 89.7] | 80.5 [70.5, 88.0] | 81.0 [73.4, 87.5] |
| Average | 90.1 ± 10 | 90.3 ± 13.8 | 89.8 ± 12.4 |
| **Prostate** | | | |
| Laterality specific cancer diagnosis (left) | 97.2 [90.8, 100.0] | 95.9 [88.6, 98.8] | 96.6 [92.2, 98.7] |
| Laterality specific cancer diagnosis (right) | 96.2 [89.4, 98.8] | 97.4 [91.7, 100.0] | 96.8 [92.5, 98.8] |
| PI-RADS | 98.2 [93.6, 99.6] | 93.3 [85.5, 100.0] | 95.1 [86.9, 100.0] |
| Average | 97.2 ± 1 | 95.5 ± 2.1 | 96.2 ± 0.9 |
| **MDS** | | | |
| Bone marrow | 93.5 [89.2, 96.5] | 97.4 [94.1, 99.0] | 95.4 [92.9, 97.2] |
| Untreated Myelodysplastic Syndrome | 91.7 [54.8, 100.0] | 68.8 [41.1, 88.9] | 78.6 [54.5, 92.9] |
| Average | 92.6 ± 1.3 | 83.1 ± 20.2 | 87 ± 11.9 |
| **Overall** | | | |
| Average | 93.9 ± 3.2 | 91 ± 5.8 | 92 ± 4.3 |

Dataset-level statistics include precision, recall, and F1 scores with 95% confidence intervals in brackets. Averages ± standard deviations per dataset are calculated across questions in each dataset, and overall averages ± standard deviations are calculated across the dataset averages.
